# Privacy-Preserving Retrieval-Augmented Generation on Local Devices for Regenerative Medicine Applications

**DOI:** 10.1101/2025.10.20.25337146

**Authors:** Tsunehiko Takamura, Akihiro Umezawa

**Author notes:** Corresponding author Akihiro Umezawa, Center for Regenerative Medicine, National Center for Child Health and Development Research Institute, 2-10-1 Okura, Setagaya, Tokyo, 157-8535, JAPAN.

## Abstract

Retrieval-augmented generation (RAG) has emerged as a promising approach to improve the factual consistency and domain-specific accuracy of large language models (LLMs), particularly in fields that demand precise and up-to-date knowledge. However, existing RAG implementations are often cloud-based and unsuitable for sensitive domains such as clinical research and regenerative medicine, where data confidentiality is paramount. In this study, we propose a privacy-preserving RAG framework using Gemma 3, a lightweight local LLM, implemented and evaluated on a commercially available MacBook Air M3. The framework operates offline without external network access, ensuring robust data security, and is feasible even in institutions without high-performance computing infrastructure. We constructed a proprietary knowledge base centered on human embryonic stem cell (ES cell)-derived hepatocyte-like cells (HAES), integrating published literature, internal consultation records, and regulatory documents. The system demonstrates context-aware generation capabilities suitable for supporting technical inquiries related to HAES applications, such as differentiation markers, safety profiles, and clinical research protocols. While the local LLM inevitably shows some limitations compared to cloud-based large models in terms of general linguistic performance, the integration of a domain-specific retrieval system substantially compensates for this gap. This work highlights the feasibility of local-device RAG frameworks in advancing sensitive biomedical applications, offering a scalable, privacy-preserving, and clinically deployable alternative to cloud-based solutions.

## Background

The application of AI in sensitive biomedical domains such as regenerative medicine presents unique challenges, particularly where data privacy and regulatory complexity intersect (Voigt & Bussche, 2017). In contexts like clinical research and institutional consultation, where navigating complex guidelines, internal documentation, and scientific evidence is routine, traditional cloud-based language models are often unsuitable due to privacy concerns. These limitations hinder the potential of AI to support medical innovation in highly regulated and confidential environments. Recent advances in artificial intelligence (AI), especially in large language models (LLMs), have opened new possibilities to assist with research workflows and compliance tasks (OpenAI et al., 2023). One notable development is retrieval-augmented generation (RAG), a technique that combines generative AI capabilities with the factual grounding of retrieval systems (Lewis et al., 2020). By incorporating external, domain-specific knowledge bases, RAG allows for the generation of accurate, context-aware responses to technical or regulatory queries. This is particularly valuable in fields like regenerative medicine, where scientific precision, timeliness, and traceability are essential.

However, most high-performance LLMs and RAG frameworks rely on cloud infrastructure, rendering them unsuitable for institutions that handle sensitive patient data or proprietary research information (Zeng et al., 2024). Local deployment is essential for privacy and institutional compliance, yet it introduces new challenges, particularly in achieving sufficient model performance on resource-constrained hardware (Paramanayakam et al., 2024; Yao et al., 2025).Lightweight models often lack the capacity of their cloud-based counterparts, and achieving real-time, relevant responses requires architectural innovations and targeted optimization (Ofer et al., 2025; Pareja et al., 2024; Yang et al., 2025). To address these challenges, we propose a practical and privacy-preserving framework that integrates a lightweight local LLM (Gemma 3) with a retrieval-augmented generation pipeline, operating entirely offline on a commercially available laptop (MacBook Air M3). This system is designed to function within security-sensitive, low-resource environments, enabling knowledge-intensive support without external GPU acceleration or internet connectivity. As a use case, we apply the system to the domain of human ES cell-derived hepatocyte-like cells (HAES) (Umezawa et al., 2025), a technically complex area within regenerative medicine. A proprietary knowledge base—compiled from scientific publications, regulatory guidelines, and internal consultation records—was constructed to power the retrieval module. We demonstrate that this architecture can generate context-specific responses suitable for supporting technical inquiries in clinical research settings.

While this paper focuses on regenerative medicine, the underlying architecture is applicable to other domains that demand secure, domain-specific AI deployment. Our primary aim is not to outperform cloud-based models in general capabilities, but to offer a deployable, auditable, and secure solution for institutions operating under strict data protection requirements. Through this work, we demonstrate that effective AI support for biomedical applications can be achieved even with modest computational resources, provided that system design is carefully aligned with privacy and domain constraints.

## Methods

### System Overview

We developed a local RAG system designed to operate entirely offline on a consumer-grade laptop. The objective was to propose a practical framework that integrates a lightweight local LLM with a retrieval module, enabling privacy-preserving, context-aware support for domain-specific queries in regenerative medicine. Figure 1 illustrates the architecture of the proposed framework.

**Figure 1.**
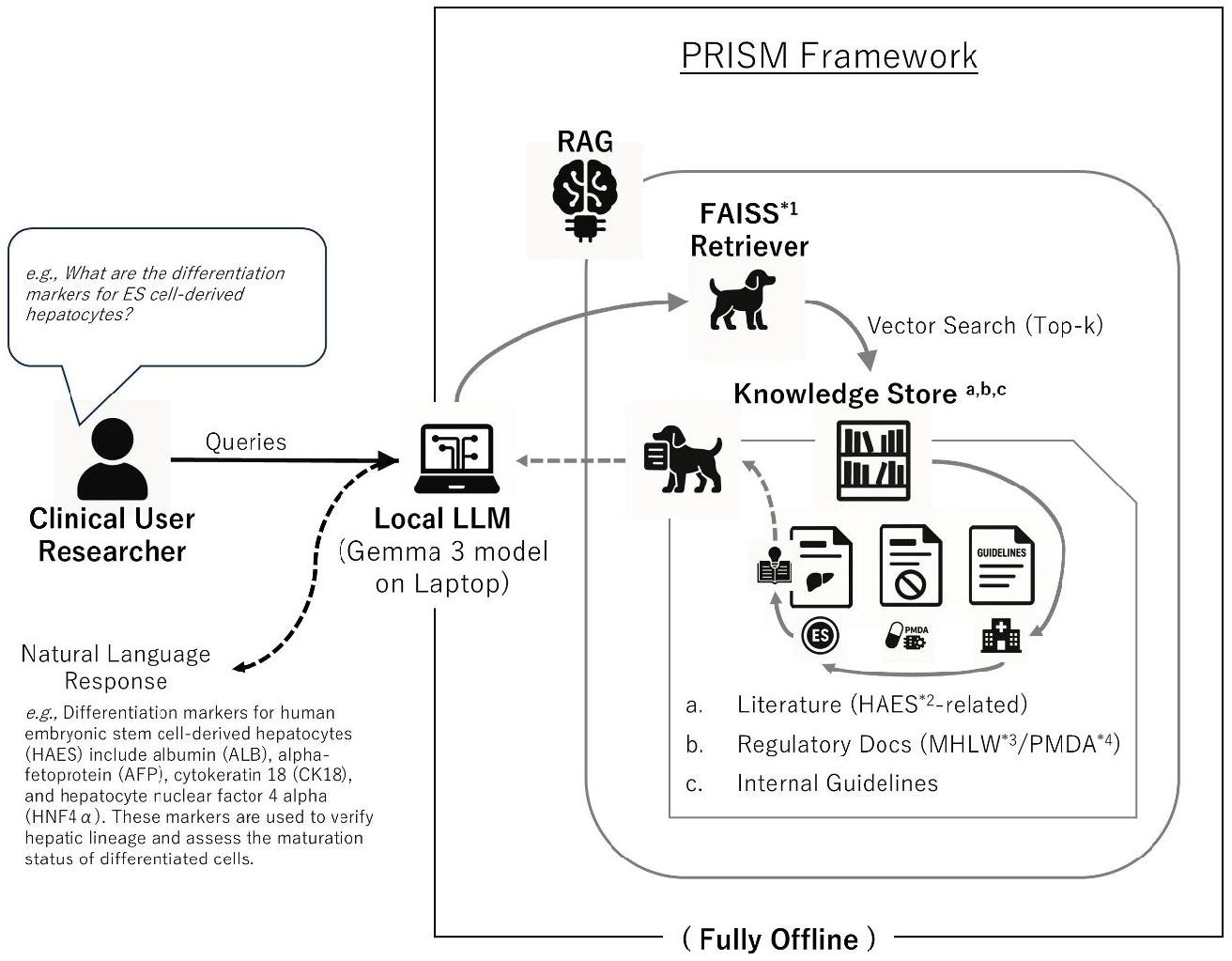
Architecture of the PRISM Framework: Privacy-preserving Retrieval for Institutional Science and Medicine. This figure illustrates the architecture of the Local RAG model used in the PRISM framework, which ensures secure and local access to scientific and medical knowledge without external data transmission. The system comprises multiple components designed for secure and domain-specific RAG. A clinical user or researcher initiates a query via the user interface. The query is processed by the retrieval module, which leverages local FAISS indexing with sentence embeddings to search a domain-specific knowledge base. This base includes: a peer-reviewed scientific literature (HAES-related), b regulatory documents (e.g., MHLW, PMDA), and c internal consultation records and institutional guidelines. The retrieved content is then passed to a local Large Language Model (Gemma 3), running fully offline on a MacBook Air M3 via LM Studio. The LLM integrates query and knowledge to produce a tailored response, which is returned to the user. All operations are executed locally, ensuring full data privacy and offline functionality in clinical and research environments. The workflow consists of the following components: · Clinical User Researcher: The process begins with a user (e.g., clinician or researcher) submitting a query through a user interface. · Local LLM (Query Input): The interface collects and formats the user query for further processing. · Retrieval Module: The query is passed to a retrieval module, where a local FAISS^*1^ (Facebook AI Similarity Search) index, combined with sentence-transformer embeddings, searches for semantically relevant information. · Knowledge Store: The retrieval module accesses a domain-specific knowledge base containing three categories of documents. a Peer-reviewed scientific literature on HAES^*2^ (Human Embryonic Stem cell-derived Hepatocytesedical Devices Agency) b Regulatory documents from agencies (e.g., MHLW^*3^; Ministry of Health, Labour and Welfare, FDA^*4^; Pharmaceuticals and Medical Devices Agency) c Internal consultation records and institutional guidelines · Local LLM (Gemma 3 on Macbook Air M3): The retrieved knowledge chunks are sent to a lightweight local large language model (Gemma 3), deployed fully offline on a consumer-grade laptop (MacBook Air M3), using LM Studio as the inference engine. The LLM integrates the query and the retrieved information to generate a context-aware response. · Response (User Output): The response is returned to the user via the interface, completing the consultation loop. This closed-loop design ensures that all operations are conducted locally, without any external network access, providing maximum data privacy and security while supporting knowledge-intensive clinical or research inquiries.

The system consists of four core components: (1) a user interface for interaction, (2) a retrieval module based on vector similarity, (3) a knowledge base constructed from validated biomedical literature and datasets, and (4) a lightweight local LLM (Gemma 3) running on an edge device (Figure 1). This design was intentionally chosen to minimize privacy risks while maintaining practical usability in clinical research environments. To highlight the practical advantages of our system over conventional approaches, Table 1 compares the key components of the PRISM framework with typical cloud-based generative AI solutions.

**Table 1.** Component-wise Comparison between the PRISM Framework and Cloud-Based GenAI Solutions.

| Component | PRISM Framework | Cloud-based GenAI |
| --- | --- | --- |
| Interface | Local UI | Web Browser / API |
| Retriever | FAISS + Document Vector Search | None |
| Knowledge | HAES-Specific Local DB | General-Purpose Pretrained Models |
| LLM | Gemma 3 (On-device) | GPT-4 / Claude (Cloud) |
| Output | Offline Instant Response | Cloud-based Response |
| Data Leakage | Fully Local Protection | Risk of External Transmission |

This table outlines the differences in system interface, retrieval method, knowledge base, LLM deployment, response modality, and data leakage risk between the proposed local framework and standard cloud-based generative AI services.

### Local LLM Setup

The language model utilized in this study is Gemma 3 (12B parameters), selected for its balance between performance and resource efficiency, allowing deployment on widely available hardware. The model was executed on a MacBook Air M3 (8-core CPU, 8GB RAM) without external GPU acceleration. Local inference was enabled via the LM Studio platform, and all operations were conducted in a fully offline environment to maximize data security.

### Knowledge Base Construction

A proprietary domain-specific knowledge base was created to support the retrieval module, comprising:

- Peer-reviewed publications related to ES cell-derived hepatocyte-like cells (HAES)
- Regulatory documents from Japanese and international agencies (e.g., MHLW, FDA)
- Internal consultation records and institutional guidelines (non-public)

All documents were processed through text chunking (maximum 512 tokens per chunk) and embedded using the all-MiniLM-L6-v2 sentence-transformer model. The resulting embeddings were stored in a local FAISS index for efficient semantic retrieval.

### RAG Pipeline

The RAG pipeline was implemented using the Dify platform, which provides integrated support for retrieval-augmented generation workflows. A local LLM was connected via an internal HTTP API, and semantic search was powered by a FAISS index of embedded document chunks. The system workflow was structured as follows:

- A user submits a query to the system.
- The retriever identifies the top-k relevant chunks (k = 20) from the FAISS index.
- Retrieved chunks are assembled into a prompt and sent to the local LLM via Dify’s orchestration layer.
- The LLM generates a context-aware response, which is returned to the user.

To accommodate local hardware constraints, the input prompt was limited to 2048 tokens.Prompt formatting and compression strategies were employed to ensure the quality and efficiency of generation.

### Evaluation Framework

To assess the performance of a local LLM integrated with a RAG pipeline, we conducted a comparative evaluation against leading cloud-based LLMs (ChatGPT, Claude, and Gemini). All models were given an identical task: to generate 10 question–answer (Q&A) pairs based on the content of a given academic PDF. In the local LLM + RAG setup, the PDF was indexed using a vector store (e.g., FAISS), and relevant document chunks were retrieved via semantic search to support answer generation. The overall evaluation pipeline, including the configuration of the local LLM with RAG and the baseline cloud-based models, is illustrated in Figure 2 and Supplemental Methods.

**Figure 2.**
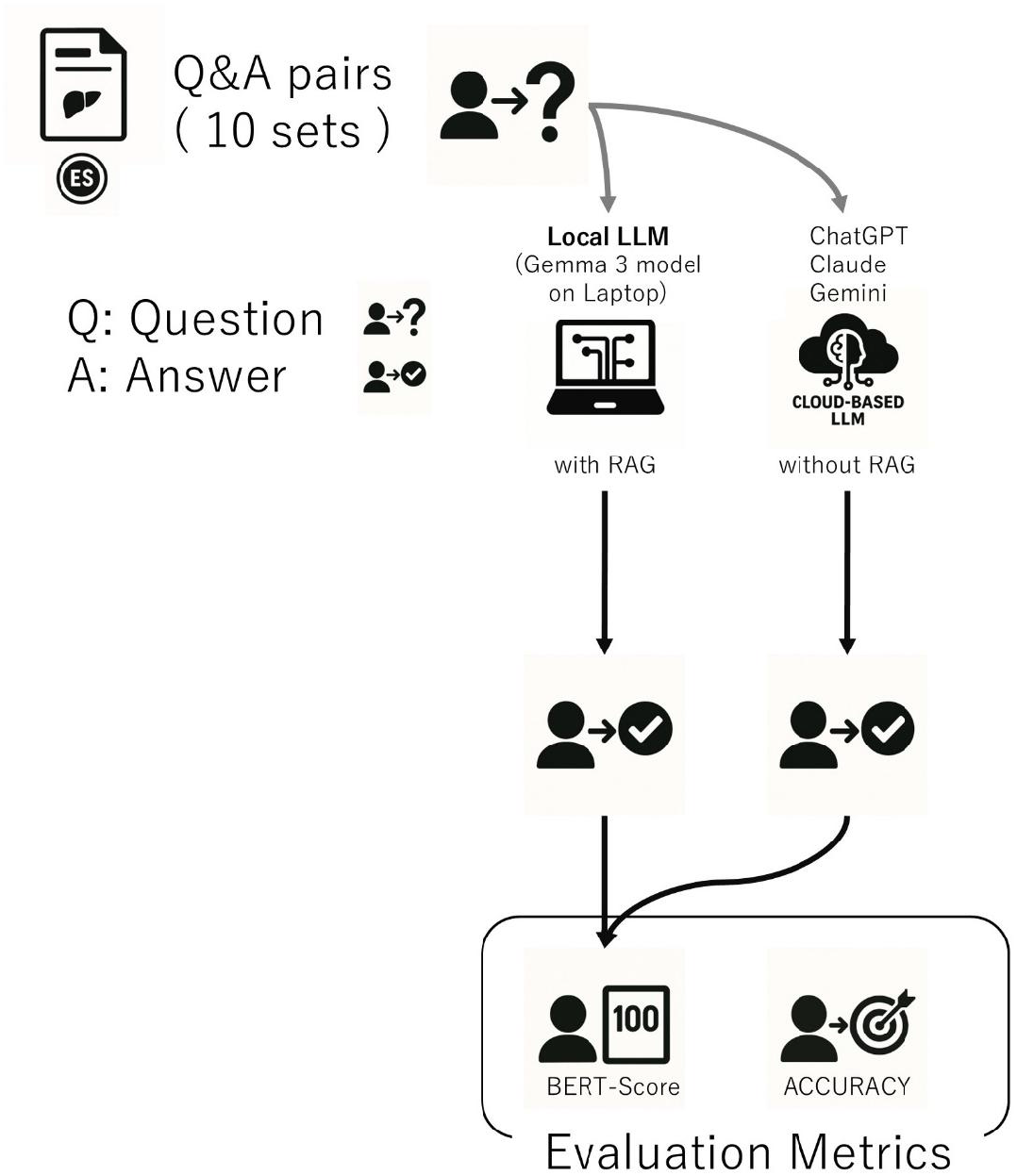
Comparative Evaluation Framework for Local LLM (Gemma 3 with RAG) vs. Cloud-Based LLMs. This figure illustrates the evaluation pipeline comparing a locally deployed LLM (Gemma 3 running on a laptop with Retrieval-Augmented Generation, RAG) against leading cloud-based LLMs (ChatGPT, Claude, and Gemini), which generate responses without RAG. All models were tasked with generating ten Q&A pairs from the same academic PDF. For the local system, the PDF was indexed in a vector store (e.g., FAISS) to enable semantic retrieval for grounding responses.Outputs from both local and cloud systems were evaluated using BERTScore for semantic similarity and binary accuracy for factual correctness, providing a reproducible framework under realistic usage conditions.

All cloud-based models were used under default configurations without any system-level prompt tuning or parameter modifications, reflecting typical user scenarios and ensuring fairness and reproducibility.

In addition to semantic similarity metrics such as BERTScore (Zhang et al., 2019), we performed a binary accuracy evaluation to assess whether each response generated by the local LLM appropriately addressed the corresponding question and aligned with the expected reference answer. Responses were labeled as either “correct” or “incorrect” based on predefined criteria emphasizing factual consistency and relevance.

It is important to note that this evaluation was not intended as a rigorous benchmark of model performance. Rather, our aim was to assess how closely a local LLM with a RAG pipeline could approximate the answer quality of state-of-the-art cloud-based language models in a practical application context.

## Results

### Use Case Simulation

Rather than performing formal benchmarking, we conducted use case demonstrations simulating realistic consultation scenarios in regenerative medicine. Representative queries are summarized in Table 2.

**Table 2.** Representative queries and example AI-generated responses in the HAES consultation domain.

| **Query** | **Example AI-Generated Response** |
| --- | --- |
| *What are the differentiation markers for ES cell-derived hepatocytes?* | Differentiation markers for human embryonic stem cell-derived hepatocytes (HAES) include albumin (ALB), alpha-fetoprotein (AFP), cytokeratin 18 (CK18), and hepatocyte nuclear factor 4 alpha (HNF4α). These markers are used to verify hepatic lineage and assess the maturation status of differentiated cells. |
| *What ethical considerations apply when using HAES in pediatric clinical trials?* | The use of HAES in pediatric clinical trials requires strict adherence to ethical standards, including obtaining informed consent from parents or legal guardians, minimizing potential risks to child participants, and ensuring oversight by institutional review boards (IRBs). Long-term monitoring of cell engraftment and immune responses is also recommended. |
| *Which regulatory documents govern the use of human ES cells in Japan?* | In Japan, the use of human ES cells is regulated under the *Guidelines on the Derivation and Utilization of Human Embryonic Stem Cells* issued by the Ministry of Health, Labour and Welfare (MHLW). Clinical research must also comply with the *Act on the Safety of Regenerative Medicine*. |

### System Performance and Responsiveness

The proposed local RAG framework was successfully deployed and operated on a MacBook Air M3 without requiring external GPU resources or cloud services. Across typical queries, average response times ranged between 3.1 and 5.7 seconds, depending on prompt length and retrieval volume. The system maintained stable performance throughout extended test sessions (n = 50 queries across 5 test sessions), demonstrating the practical feasibility of lightweight local deployment for domain-specific knowledge support in regenerative medicine.

### Impact of RAG Integration

To assess the qualitative benefits of retrieval augmentation, system outputs were compared between two modes:

- Gemma 3 is operating without retrieval support, and
- Gemma 3 is operating with RAG integration.

A total of 30 Q&A items were systematically evaluated under two configurations of the Gemma 3 model: one without any retrieval support, and the other incorporating a RAG mechanism. For each question, the locally generated response was directly compared to reference answers previously produced by state-of-the-art cloud-based large language models (ChatGPT, Claude, and Gemini), which served as proxies for high-quality, expert-level outputs. Two primary evaluation metrics were used to assess performance: semantic similarity using BERTScore (F1) (Zhang et al., 2019), and binary accuracy based on correctness (Figure 3, Supplemental Data).

**Figure 3.**
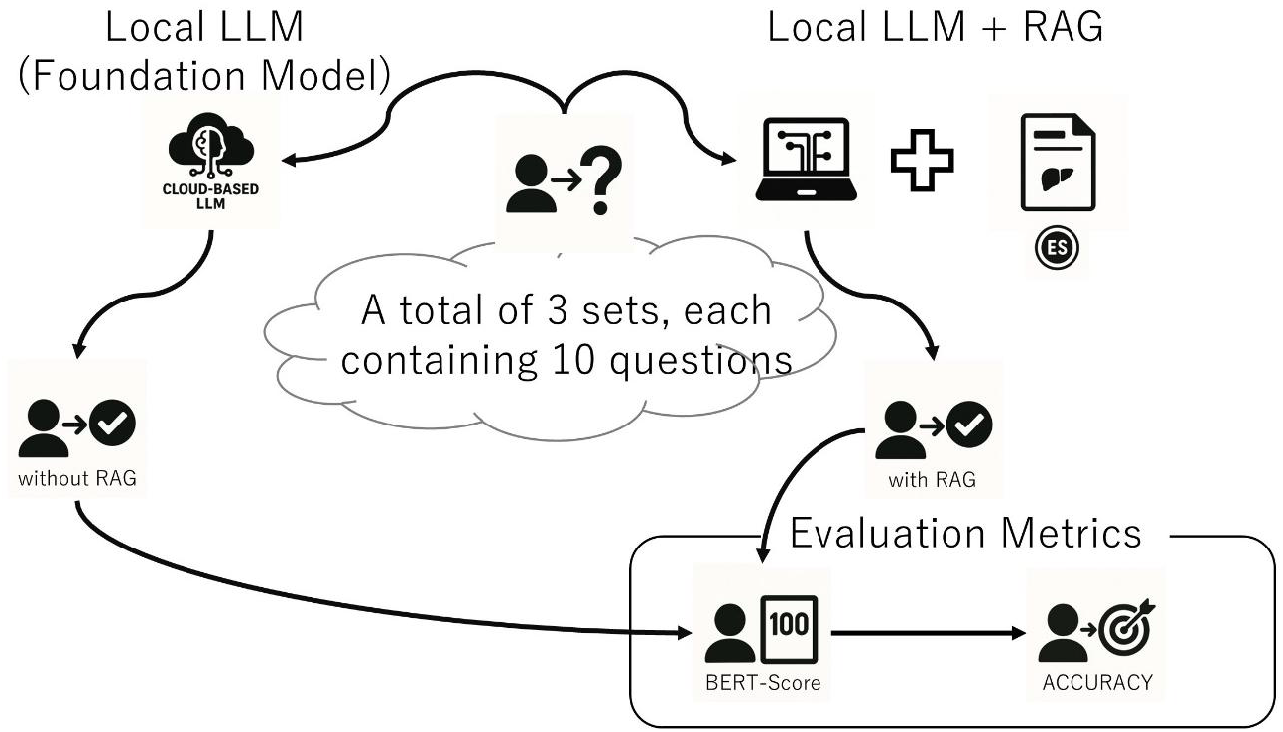
Impact of RAG Integration on Local LLM Performance. To assess the qualitative benefits of retrieval augmentation, system outputs were compared between two configurations of the Gemma 3 model: one operating without retrieval support, and another integrated with a RAG mechanism. A total of 30 Q&A items were evaluated in both settings. For each item, responses generated by the local model were compared against reference answers provided by state-of-the-art cloud-based LLMs (ChatGPT, Claude, Gemini). Evaluation metrics included semantic similarity using BERTScore (F1) and binary accuracy based on correctness.

The semantic similarity between locally generated responses and cloud-based reference answers, as quantified by BERTScore (F1) (Zhang et al., 2019), remained largely comparable across the two configurations. Specifically, the non-RAG setting yielded a BERTScore of 0.885, while the RAG-integrated system achieved a score of 0.883. These nearly identical values indicate that, at the level of surface-level semantic overlap, both configurations generated outputs that were similarly aligned with the target responses in terms of language and meaning.

In contrast, binary accuracy—defined as whether each answer directly and appropriately addressed the given question while remaining factually consistent with the reference answer— revealed a more substantial difference. The RAG-enabled configuration demonstrated a significant improvement in correctness, achieving an accuracy of 86.7% (26 out of 30 correct), whereas the non-RAG configuration achieved only 60.0% (18 out of 30 correct). This suggests that while lexical similarity alone did not differ markedly, the inclusion of retrieval-based context significantly enhanced the factual reliability and task-specific relevance of the model’s outputs.

These findings underscore the role of retrieval augmentation not only in enriching the factual basis of responses but also in bridging the gap between generic language modeling and domain-specific knowledge application.

Importantly, all evaluations were performed under strictly identical prompt conditions, without the use of model-specific instruction tuning, manual annotation, or auxiliary context injection, ensuring that observed differences were attributable solely to the presence or absence of RAG.

The RAG-integrated configuration consistently yielded higher-quality outputs across all evaluated criteria, providing more contextually accurate, factually grounded, and semantically aligned answers. These results demonstrate that even in a resource-constrained local environment, without the need for cloud access or external GPU acceleration, integrating RAG can meaningfully enhance the domain specificity, functional utility, and overall reliability of AI-generated content in specialized fields such as regenerative medicine.

### Practical Utility in Knowledge-Intensive Tasks

In simulated consultation scenarios involving clinical trial preparation, regulatory documentation drafting, and the interpretation of internal guidelines, the system demonstrated potential utility in supporting technical inquiry workflows. Although a formal user study was not conducted, preliminary impressions from a domain expert indicated that the system may contribute to reducing manual document retrieval efforts and offer satisfactory assistance during early-stage information gathering in specialized biomedical contexts.

While human oversight remains indispensable for complex interpretive tasks, the proposed framework functioned effectively as an auxiliary tool for preliminary research, documentation support, and knowledge navigation within sensitive and information-dense biomedical domains.

## Discussion

The framework proposed in this study—integrating a lightweight local LLM with RAG— demonstrated practical viability in highly specialized domains such as regenerative medicine, where both domain specificity and strict data confidentiality are critical. In use cases such as regulatory consultation and clinical trial preparation, the system effectively retrieved and synthesized contextually relevant information from a domain-specific knowledge base comprising clinical guidelines, regulatory documents, and technical literature. This capacity suggests that the framework can streamline documentation review and preliminary decision-making, enhancing the speed and relevance of expert responses during early research and development stages.

A particularly meaningful aspect of this work is that the entire system operates on a commercially available laptop under constrained computational resources, without reliance on external GPUs or cloud infrastructure. This design enables deployment in environments with limited IT capacity and strict privacy requirements. The tailored knowledge base and RAG pipeline significantly compensated for the intrinsic limitations of lightweight models, demonstrating a viable strategy for balancing utility and security.

In addition to improved response accuracy, the system also appeared to enhance workflow efficiency in early-stage technical consultations. These findings suggest that AI systems like ours could evolve beyond passive information providers and serve as active cognitive partners for domain experts navigating complex regulatory and clinical knowledge.

While lightweight local LLMs alone may have limited capacity for high-quality response generation, combining them with RAG allows for performance approaching that of general-purpose cloud-based models. The proposed PRISM framework thus emerges as a practical, secure solution for institutions handling sensitive, domain-specific tasks.

Moreover, the local deployment of PRISM offers key advantages over cloud-based alternatives by enabling full on-device processing and reducing dependency on external services. The ability to perform full on-device processing is particularly valuable in healthcare and life sciences, where stringent requirements for data privacy, auditability, and regulatory compliance apply. Although the current implementation uses lightweight LLMs that inherently have some limitations (Hinton et al., 2015; Wang et al., 2024), ongoing advancements in foundation model technology are expected to significantly improve their performance. As such, even if the PRISM framework currently does not fully match the capabilities of cloud-based systems, it is well-positioned to become increasingly competitive as local models mature. We anticipate that PRISM will serve as a foundational platform for the broader adoption of privacy-preserving AI systems across sensitive biomedical domains.

Despite its promise, this study has several limitations. First, part of the RAG system includes confidential information, which prevented us from evaluating those components due to the risk of data leakage. As a result, the evaluation was limited to publicly available academic literature, and for practical reasons, we focused on a single research paper. It is important to note that the aim of this study was not to benchmark model performance, but to demonstrate the feasibility of implementing a local RAG system in a privacy-preserving biomedical context.

While the integration of RAG improved output quality in our PDF-based evaluation, it is not a comprehensive solution. Its performance on complex clinical queries—such as those involving ethical ambiguity or evolving regulatory standards—was not assessed in this study. These types of queries may yield ambiguous responses without clear contextual or normative guidance, requiring particular caution in future applications (Amugongo et al., 2025; Gargari & Habibi, 2025). In such cases, human oversight remains essential to ensure interpretive accuracy and appropriate contextual judgment (Olabiyi et al., 2025).

Managing sensitive data locally continues to pose significant operational challenges, warranting particular attention. While local deployment mitigates the risk of external data leakage by avoiding transmission to third-party servers, it does not eliminate the risk of information exposure altogether. Sensitive data retained within institutional servers or local devices remains vulnerable to internal threats, such as improper access, insufficient encryption, or inadequate device-level security.Accordingly, strict data governance and comprehensive security protocols are essential to ensure end-to-end protection, even in locally managed environments.

Compared to general-purpose cloud-based LLMs such as ChatGPT, our local RAG system demonstrated superior performance in domain-specific tasks, thanks to its tailored knowledge base and privacy-preserving architecture. However, it still lacks the general reasoning capacity and cross-domain versatility of large-scale cloud models. Accordingly, the local RAG framework should be understood not as a universal replacement, but as a targeted solution for settings where cloud-based models are impractical—such as clinical institutions handling sensitive or regulated data. Several development directions have been proposed in the field to enhance system robustness and applicability, including knowledge base expansion, training on complex consultation scenarios, and integration with institutional knowledge systems (Amugongo et al., 2025; Gargari & Habibi, 2025; Leszczyński et al., 2025). Establishing local RAG systems as a standard in healthcare could transform clinical practice by enabling institutions to process sensitive patient data entirely on-site. This shift reduces reliance on external servers, minimizes data exposure risks, and facilitates compliance with evolving privacy regulations such as GDPR and HIPAA. As these regulatory frameworks continue to tighten, the demand for secure, localized AI infrastructure is expected to grow accordingly.

While further validation is required, this study presents a proof of concept and outlines a practical framework for developing domain-specific AI systems tailored to clinical contexts. Our findings suggest that such systems can be realized using modest hardware and open-source tools—without necessarily compromising privacy, scalability, or autonomy—while meeting concrete regulatory and operational requirements.

In future work, we plan to expand the PRISM framework to support additional biomedical domains and conduct formal, multi-expert evaluations to quantitatively assess its performance and operational utility.

## Data Availability

The datasets and cells used during the current study are available from the corresponding author upon reasonable request.

## Declarations

### Ethics approval and consent to participate

This study did not involve human participants, human data, or animal experiments. According to the relevant institutional and national regulations, ethics approval and consent to participate were not applicable.

### Consent for publication

Not applicable.

### Authors’ contributions

TT and AU conceived and designed the study. TT performed the data analysis. TT and AU discussed the results and interpretation. TT and AU drafted and revised the manuscript. All authors read and approved the final manuscript.

## Acknowledgments

We would like to express our sincere thanks to E. Suzuki and K. Saito for secretarial work.

## Declaration of generative AI and AI-assisted technologies in the writing process

During the preparation of this work, TT and AU used ChatGPT, Perplexity AI, and Google Translate to assist with English writing. These tools were used solely for language editing and phrasing support, not for generating scientific content or analysis. After using these tools, TT and AU carefully reviewed and edited the content, and take full responsibility for the final manuscript.

