## Supplemental Methods for "Privacy-Preserving Retrieval-Augmented Generation on Local Devices for Regenerative Medicine Applications"

**Appendix A**

**Instructions for Generating Question–Answer (Q&A) Sets**

To assess comprehension of the research article, we designed a set of instructions to guide the generation of question–answer (Q&A) items derived directly from the content of the paper. These instructions were used to create a standardized evaluation framework for both cloud-based and locally deployed language models.

**Objective**

The goal of the Q&A generation process was to produce a practical and content-aligned set of questions that accurately reflect key information in the article and can be used to evaluate understanding.

**Generation Rules**

Each question must be based on explicitly stated information in the paper.

Questions should reflect what a non-expert but interested user might reasonably ask.

Answers must be accurate summaries of content present in the article; speculative or inferred content should be avoided.

Answers should be concise (1–3 sentences) while preserving factual precision.

Whenever applicable, indicate the section of the paper (e.g., Introduction, Methods, Results, Discussion) to which the Q&A item pertains.

**Output Format**

Each Q&A pair should follow the format below:

Q: (The question)

A: (The corresponding answer)

Section: (Relevant section of the article)

**Appendix B**

**Prompt Template for Local LLM + RAG Evaluation**

To evaluate the ability of the local LLM integrated with a retrieval-augmented generation (RAG) pipeline, the following prompt was used to assess factual alignment and response accuracy against reference questions derived from the academic paper.

**Prompt Instruction (English Translation)**

You are a reading assistant for a scientific research paper. Based on the retrieved contextual information already provided through the RAG mechanism, please answer the following questions:

[Instructions]

Answer each question based solely on the retrieved information.

If no explicit information is found, respond with:

"Information not found."

Keep each answer concise (1–3 sentences).

[Questions]

What is the main objective of this study?

What was the target body weight set for safe liver transplantation?

What were the key safety findings observed in preclinical testing of ESC-derived HLCs?

Through which route were HLCs administered in the clinical trial?

How many neonates participated in the clinical trial?

What clinical outcomes were achieved in all patients following HLC infusion?

What was the most concerning complication observed after human HLC administration?

What was the differentiation efficiency of HLCs?

Was there any evidence of long-term retention of HLCs in the liver?

What methods are planned to more accurately evaluate HLC function in the future?

What was the therapeutic goal of the cell treatment used in this study?

How many patients were enrolled in the clinical trial, and what disorders did they have?

How was the administration of HLCs carried out?

What animal models were used in the preclinical studies?

Did all patients reach the target body weight?

What major complications were observed following HLC treatment?

What notable properties of HLCs were confirmed?

How was the safety regarding tumorigenicity assessed?

What immunosuppressive agents were used?

What was the final conclusion of the study?

What are the symptoms of urea cycle disorders (UCD)?

Why is liver transplantation not immediately performed in neonates with UCD, despite being an effective treatment?

How many neonates received HLC infusion therapy during the clinical trial?

How was the safety of HLCs evaluated in preclinical trials?

How were HLC infusions administered?

What functions did HLCs demonstrate in vitro?

What important body weight target was achieved by all patients prior to liver transplantation?

How did patients' ammonia levels change after HLC treatment?

What aspects of long-term safety will be evaluated in follow-up studies on HLC therapy?

What is the primary purpose of this study?
